# Differential effects of theta-gamma tACS on motor skill acquisition in young individuals and stroke survivors: a double-blind, randomized, sham-controlled study

**DOI:** 10.1101/2024.06.10.24308694

**Authors:** L.S. Grigutsch, B. Haverland, L. S. Timmsen, L. Asmussen, H. Braaß, S. Wolf, T. V. Luu, C.J. Stagg, R. Schulz, F. Quandt, B.C. Schwab

**Author notes:** Corresponding authors: Bettina C. Schwab, University of Twente, Drienerlolaan 5, 7522 NB Enschede, The Netherlands,; Fanny Quandt, University Medical Center Hamburg-Eppendorf, Martinistraße 52, 20246 Hamburg, Germany. shared last authorship (F. Quandt, B.C. Schwab).

## Abstract

**Background:** Theta-gamma transcranial alternating current stimulation (tACS) was recently found to enhance thumb acceleration in young, healthy participants, suggesting a potential role in facilitating motor skill acquisition. Given the relevance of motor skill acquisition in stroke rehabilitation, theta-gamma tACS may hold potential for treating stroke survivors.

**Objective:** We aimed to examine the effects of theta-gamma tACS on motor skill acquisition in young, healthy participants and stroke survivors.

**Methods:** In a pre-registered, double-blind, randomized, sham-controlled study, 78 young, healthy participants received either theta-gamma peak-coupled (TGP) tACS, theta-gamma trough-coupled (TGT) tACS or sham stimulation. 20 individuals with a chronic stroke received either TGP or sham. TACS was applied over motor cortical areas while participants performed an acceleration-dependent thumb movement task. Stroke survivors were characterized using standardized testing, with a subgroup receiving additional anatomical brain imaging.

**Results:** Neither TGP nor TGT tACS significantly modified general motor skill acquisition in the young, healthy cohort. In contrast, in the stroke cohort, TGP diminished motor skill acquisition compared to sham. Exploratory analyses revealed that, independent of general motor skill acquisition, healthy participants receiving TGP or TGT exhibited greater peak thumb acceleration than those receiving sham.

**Conclusion:** Although theta-gamma tACS increased thumb acceleration in young, healthy participants, consistent with previous reports, it did not enhance overall motor skill acquisition in a more complex motor task. Furthermore, it even had detrimental effects on motor skill acquisition in stroke survivors.

## Introduction

Transcranial alternating current stimulation (tACS) applies weak electric currents to the scalp. It has the potential to modulate neural activity noninvasively [1–3], with clear behavioral effects [4–6]. Given its relatively easy application and high tolerability, tACS may be ideally suited for clinical applications. With the idea of modifying pathological oscillatory activity, tACS has been suggested as a potential future treatment for several neurological and psychiatric diseases, including Parkinson’s disease [7–9], schizophrenia [10,11], and obsessive-compulsive disorder [12,13].

In stroke survivors, tACS has been investigated as a tool to modulate neural activity and connectivity [14–17], a promising approach as oscillatory activity has been shown to change after a stroke [18,19]. Nevertheless, no study has aimed to improve hand-motor skill acquisition after stroke with tACS. Motor skill acquisition holds a pivotal role in stroke rehabilitation as stroke survivors have to re-acquire motor skills with their affected limbs.

Theta-gamma tACS, which combines theta (6 Hz) and gamma (75 Hz) rhythms into a single waveform, has been shown to improve motor skill acquisition in healthy participants. Notably, motor skill acquisition is only improved when coupling gamma oscillations to the peak of the theta wave (TGP) compared to coupling gamma to the theta trough (TGT) or using sham stimulation [4]. Consequently, this specific form of tACS presents a promising approach to be tested in stroke survivors with motor impairments.

High-gamma oscillations (60-100 Hz) are time-locked to movement onset [20] and are hypothesized to represent a movement execution signal [21,22]. In the neocortex, the amplitude of fast oscillations is frequently modulated by the phase of slower oscillations (phase-amplitude coupling, PAC). PAC between theta and gamma frequencies in the rodent hippocampus and entorhinal cortex is associated with exploratory behavior, learning, and memory processes [23–25]. Theta-gamma PAC has also been observed in humans [26], primarily associated with hippocampal learning, long-term- and working memory and cognitive control [27–30]. Further, the success of motor learning has been reported to increase with the amount of theta-gamma PAC in the motor cortex [31]. Also, a recent study in stroke survivors demonstrated that the amount of theta-gamma PAC in the primary motor cortex (M1) correlates positively with motor recovery throughout rehabilitation [32]. In sum, theta-gamma PAC in motor cortical areas could be relevant for motor skill acquisition.

Here, we hypothesized that in an acceleration-dependent thumb movement task, TGP tACS would improve motor skill acquisition compared to TGT tACS and sham. First, we aimed to confirm this hypothesis in 78 young, healthy volunteers. Second, we investigated the effects of theta-gamma tACS in 20 chronic stroke survivors with the idea of a potential future use in stroke rehabilitation.

## Materials and methods

### Participants and study protocol

#### Young cohort

78 right-handed adults between 18 and 35 years successfully completed the experimental session. The following exclusion criteria were applied: history of neurological or major psychiatric illness, pronounced cognitive deficits, regular intake of psychotropic medication, pregnancy, and exclusion criteria for tACS (history of severe head trauma or brain surgery, devices or implants in the head region, implanted electric devices, epilepsy or history of an epileptic seizure). In total, 84 participants were recruited from the local community and participated in the study. Six participants had to be excluded (three due to technical problems, two due to pain caused by tACS, and one due to committing errors in > 25% of trials).

#### Stroke cohort

20 individuals with a stroke, confirmed by imaging, in the chronic phase (at least six months after stroke) were recruited. They had had no prior clinical stroke and had experienced an initial hand-motor impairment lasting at least 24 hours. The following exclusion criteria were pre-registered: history of major psychiatric illness or neurological disease other than stroke, pronounced cognitive deficits, regular intake of psychotropic medication, pregnancy, and exclusion criteria for tACS. After the pre-registration, minor changes to the exclusion criteria were made, and candidates were not excluded if (i) taking low doses of medication for neuropathic pain or (ii) suffering from a neurological disease not affecting the brain or the performing hand. In total, 23 stroke survivors participated, but three had to be excluded retrospectively (two due to errors in the experiment, one due to intake of psychotropic medication). All stroke survivors were characterized using standardized testing of global disability and motor function: modified Rankin Scale (mRS), National Institutes of Health Stroke Scale (NIHSS), Mini-Mental Status Test (MMST), Edinburgh Handedness Inventory (EHI), Upper Extremity Fugl-Meyer-Assessment (UEFM), Action Research Arm Test (ARAT), Nine Hole Peg Test (NHPT), Box and Block Test (BBT) and maximal grip strength. Stroke survivors eligible for magnetic resonance imaging (MRI) received structural brain imaging.

#### Study Protocol

The study part on young, healthy participants was pre-registered on the Open Science Framework (OSF) platform (https://osf.io/mqwt5), and the study part on stroke survivors was pre-registered on the platform clinicaltrials.gov (Identifier: NCT05576129). The study was approved on June 7th, 2021, by the local ethics committee of the Medical Association of Hamburg (2021-10410-BO-ff) and conducted in accordance with the Declaration of Helsinki. Participants gave written informed consent.

#### Randomization and Blinding

All participants and those researchers interacting with participants or involved in data analysis were blinded to the group assignment until the analysis of primary outcomes was completed. The young cohort was pseudo-randomized to either TGP, TGT, or sham stimulation, equally distributed for sex. Participants in the stroke cohort were assigned to either TGP or sham stimulation, balanced for age, lesioned hemisphere, handedness, and dexterity (NHPT result of the affected hand).

### Thumb movement task

Participants of both experiments received tACS while performing a thumb abduction-adduction movement task (Figure 1A). Upon receiving a visual cue, they alternately pressed the red and the green buttons on a button box in the order red-green-red-green. The right-handed young cohort used their left thumb, whereas stroke survivors used the thumb on their stroke-affected side. The arm was immobilized in a fixture to ensure isolated thumb movements. Participants were first given five trials to familiarize themselves with the task and then instructed to complete the movement as fast as possible in all future trials. They performed 20 baseline trials. Then, tACS was started, and they performed another six blocks consisting of 40 trials each. The button sequence remained the same during the whole session and for all participants. The time required to complete an entire sequence served as the performance measure movement duration. Participants were encouraged to reduce their movement duration continuously and received visual online feedback in the post-baseline blocks. Only trials with the correct button sequence, started within one second after the Go-signal and finished within a maximum of four seconds, were considered valid. A 3D acceleration sensor (Brain Products GmbH, Gilching, Germany) was fixed to the tip of the thumb, and acceleration was recorded in three dimensions of space for exploratory analysis using PyCorder. The task was programmed in MATLAB version 2020b with Psychtoolbox [33].

**Figure 1:**
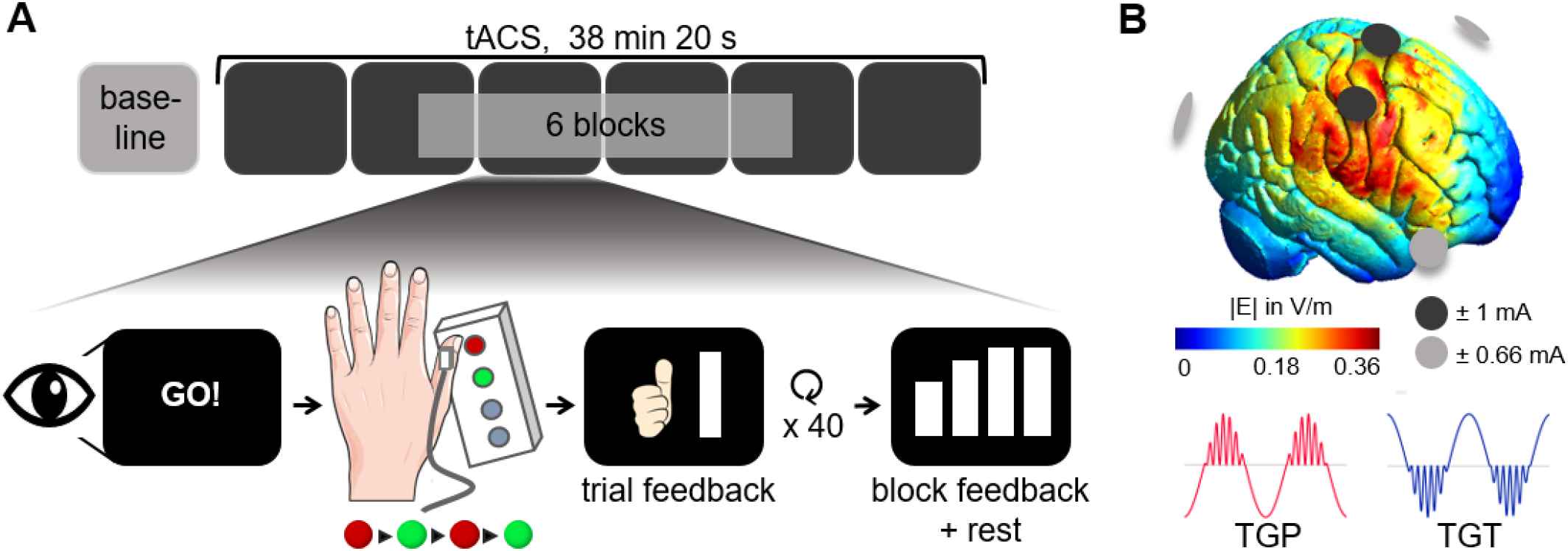
Experimental design. (A) Motor skill acquisition task: Participants performed a thumb abduction-adduction movement, pressing the buttons on a button box in the order red-green-red-green as fast as possible. They received feedback on their movement duration after each trial with a thumbs-up and thumbs-down symbol indicating whether they had improved or worsened, respectively. After each block, additional feedback on the block mean of movement duration was given. (B) tACS setup: Top: Simulation of the electric field of tACS covering the right M1 using five electrodes. Below: tACS waveforms, 6 Hz theta rhythm with 75 Hz gamma waves coupled either to the theta peak (TGP) or trough (TGT).

### tACS

HD-tACS was administered with a Starstim 8 stimulation device (Neuroelectrics, Barcelona, Spain). We conducted pilot experiments to determine the best electrode positions and current intensities to achieve high electric field strength over the motor and premotor cortices while keeping sensory side effects tolerable in all participants. We targeted the motor cortex contralateral to the performing hand, ipsilesional in the stroke cohort. Two central electrodes were placed over M1, at C2 and C4 when stimulating the right hemisphere and at C1 and C3 when stimulating the left hemisphere according to the international 10-20 system. Three return electrodes were positioned at F8, Oz, and FC1 or F7, Oz, and FC2, respectively. We used round Pistim Ag/AgCl electrodes with a contact area of 3.14 cm^2^ and conductive gel. Impedance was brought to values below 10 kΩ in all electrodes before starting the stimulation session and did not exceed 20 kΩ thereafter. The maximum current intensity was 1 mA peak-to-baseline at each of the two central electrodes and 0.67 mA at each of the three return electrodes, thus reaching a total current of 2 mA peak-to-baseline. To reduce sensations on the scalp, a local anesthetic cream was applied to the skin at the electrode positions one hour prior to stimulation.

Participants received one out of three different tACS conditions (Figure 1B, lower panel):

i. Active condition – theta-gamma peak-coupled tACS (TGP): 75 Hz gamma waves coupled to the peak of 6 Hz theta waves.
ii. Active control condition (only in the young cohort) – theta-gamma trough-coupled tACS (TGT): 75 Hz gamma waves coupled to the trough of 6 Hz theta waves.
iii. Control condition - sham stimulation: 10-second TGP stimulation at the beginning of each of the six blocks.

Each condition included a 3-second ramp-up and ramp-down period. The total duration of stimulation was 38 min 20 s in the active conditions and 60 s in the sham group.

### Electric field simulations

Electric fields of tACS were simulated using Complete Head Anatomy Reconstruction Method models in SimNIBS [34]. We defined the peak E-field strength as the 99.9th percentile of the electric field and focality as the tissue volume receiving electric field strengths above the 75th percentile. The simulation of our final tACS configuration on the MNI 152 head model (Figure 1B, upper panel) indicates that on average a peak E-field strength of 0.362 V/m and a mean E-field strength in the hand knob area (MNI coordinates 38, −22, 54, radius 1 cm) of 0.255 V/m were reached. MRI data were collected from four stroke survivors in the TGP group, and the electric field was simulated on individual head models constructed from the T1 and FLAIR images (Supplementary Figure 1).

### Post-tACS questionnaires

After the tACS session, qualitative questionnaires were obtained to estimate the perception of side effects. Participants could report skin sensations on the scalp of five qualities: warmth, itching, pulsing, stinging, and pain, and rate their intensity as “absent”, “weak”, “moderate”, “pronounced”, or “intense”. The time course of sensations could be rated as “only at the beginning”, “decreasing”, “stable”, “increasing” or “only at the end”. Participants could report any perceived flickering lights (phosphenes) and their position in the visual field and rate them on the same scales. Finally, participants were asked to guess whether they had received active or sham stimulation.

For the analysis of skin sensations, an overall score for skin sensations was computed by aggregating the individual scores for all distinct sensation qualities and considering whether they occurred only at the beginning or throughout tACS. Therefore, the ratings were converted to numbers from 0 = “absent” to 4 = “strong”. The final score was leveled as follows: 0 = “no skin sensations”, 1 = “skin sensations only at the beginning”, 2 = “sum ≤ 2”, 3 = “sum ≤ 4”, 4 = “sum > 4”.

### Brain imaging & lesion location

Structural brain images were acquired of seven participants using a 3 T Prisma MRI scanner (Siemens Healthineers, Erlangen, Germany) equipped with a 64-channel head coil. T1-weighted anatomical images were obtained with a 3-dimensional magnetization-prepared rapid gradient echo sequence (repetition time (TR) = 2500 ms, echo time (TE) = 2.15 ms, flip angle 8°, 288 coronal slices with a voxel size of 0.8 × 0.8 × 0.8 mm^3^). T2-weighted images were acquired by using a fluid-attenuated inversion recovery (FLAIR) sequence (TR = 9210 ms, TE = 92 ms, inversion time (TI) = 2500 ms, flip angle 140°, 70 axial slices with a voxel size of 0.9 × 0.9 × 2.0 mm^3^). ITK-SNAP version 4.0.1 [35] was used for the delineation of stroke lesions and the calculation of the individual lesion volumes. For the lesion map, stroke lesions were registered to a Montreal National Institute (MNI) 1 mm^3^ template and right-hemispheric lesions were flipped to the left hemisphere. For participants not eligible for MRI, either MR data from previous studies, clinical imaging data, or a hospital discharge letter with information on the lesion location was available (see Supplementary Table 1).

### Data analysis

Data analysis was performed with MATLAB version R2022b [36] and the FieldTrip toolbox [37].

#### Motor skill acquisition and movement duration

The primary outcome motor skill acquisition was defined as the relative improvement in movement duration from baseline:

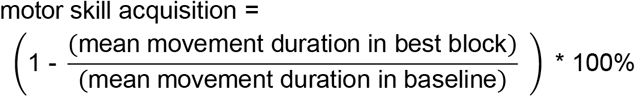

For each participant, correct trials with a movement duration more than three standard deviations away from the mean were excluded as outliers in the baseline and the six blocks, according to our pre-registered analysis plan. For each participant, the block with the lowest mean movement duration was defined as the best block. In the young cohort, participants were excluded and replaced if over 25% of baseline or post-baseline trials were missing after outlier removal, which excluded one participant.

#### Peak acceleration

Acceleration data were cut into single trials covering the movement period. Trial intervals were defined based on visual inspection (in young participants and one stroke survivor) or based on markers for the “Go” signal and the last button press in each trial (in the stroke cohort). Incorrect trials and trials with outliers of movement duration were excluded. Data were baseline-corrected on a trial-by-trial basis by subtracting the mean value. The net acceleration *a* at each time point *t* was computed as the square root of the sum of squared accelerations in each dimension of space, ax, ay, and a_z_:

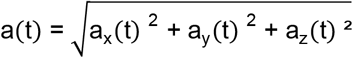. We determined the peak acceleration, defined as the maximum of a per trial. Two healthy participants were excluded from the acceleration-based analysis, as baseline acceleration data were not available.

#### Statistical Analysis

Statistical analyses were performed in R, version 4.3.2 [38] and MATLAB version R2022b [36]. Statistical significance was defined as alpha < 0.05. In each cohort, we assessed whether participants improved over the task by comparing their mean movement duration in the baseline and the last block in paired two-tailed t-tests. The number of mistakes and outliers were compared among conditions using unpaired two-tailed t-tests, Wilcoxon rank sum tests, or one-way Analyses of Variance (ANOVA), as appropriate. The primary outcome motor skill acquisition and the mean baseline movement duration were compared between stimulation groups using unpaired two-tailed t-tests or Wilcoxon rank sum tests as appropriate. We used two-tailed tests to identify the positive behavioral effects of tACS, as well as potentially relevant detrimental effects.

We hypothesized that movement duration and peak acceleration improve throughout the experiment in all groups but that there is a larger improvement in the TGP group compared to TGT and sham. To address this hypothesis, we fitted linear mixed-effects models (LME) using maximum likelihood with the lme4-package [39]. Movement duration or peak acceleration during the blocks were dependent variables. TACS stimulation condition and the time variable block and their interaction were tested as fixed effect factors. We controlled for baseline performance by including the mean movement duration or peak acceleration at baseline as a fixed effect factor. To account for interindividual differences, we included a random intercept for each ID and a random slope for the effect of block for each ID. Finally, the relationship between movement duration and peak acceleration was examined with an LME with movement duration as the dependent variable, peak acceleration as a fixed effect factor, a random intercept for each ID and a random slope for the effect of peak acceleration for each ID. P-values for fixed effects were obtained by testing the full model against the reduced model without the factor in question with the likelihood ratio test (LRT). Confidence intervals for continuous fixed effects were estimated using the profile likelihood method. In the post-hoc analysis of categorical fixed effect factors and interaction effects, we contrasted the estimated marginal means or slopes, respectively, using the emmeans-package [40] with Kenward-Roger’s method for degrees of freedom approximation. We visually inspected residual plots to detect deviations from the linear model assumptions. P-values were adjusted for multiple comparisons with Tukey’s method. The estimates for all fixed effects are reported in detail in Supplementary Table 4-5. In the exploratory analysis, we examined whether tACS effects could be biased by tACS-related skin sensations or clinical characteristics. Fisher’s exact test, Wilcoxon rank sum test, or unpaired t-tests were used as appropriate to compare these measures among groups and subgroups. The association of parameters measured on an ordinal or higher scale level with motor skill acquisition or peak acceleration improvement was tested with Kendall’s and Spearman’s correlation. For this analysis, peak acceleration improvement was defined analogous to motor skill acquisition as

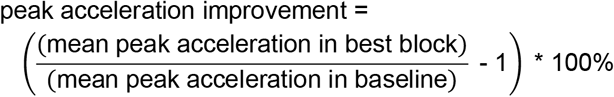

with “best block” being the block with the highest mean peak acceleration. For parameters on a nominal scale, specifically lesion location and sex, their effect on the tACS effect was examined as follows: The tACS groups were matched for the parameters in question by leaving out participants, and the LME was re-calculated with all possible participant combinations, testing for the condition main effect and the condition x block interaction effect.

## Results

### Participants

78 right-handed young adults (mean age 24.6 years, range 18-35 years, 36 male) and 20 individuals with a chronic stroke (mean age 65.2 years, range 40-83 years, 17 male) successfully completed the experiment.

focality varied between 2.5 ml and 11.7 ml.

### Clinical characteristics and structural imaging of stroke survivors

Stroke survivors showed mild to moderate upper extremity motor impairment (Supplementary Table 1, median UEFM 60). Lesions were located in subcortical and cortical brain regions (for the lesion map, see Figure 2), with a median lesion volume of 10.8 cm^3^ in those participants with available MRI. Importantly, clinical characteristics did not differ between the TGP and sham group (Table 1). In those stroke survivors who received active tACS and an MRI was obtained (n = 4), simulated peak E-field strengths ranged between 0.25 V/m and 0.42 V/m (Supplementary Figure 1), and focality varied between 2.5 ml and 11.7 ml.

**Table 1:**
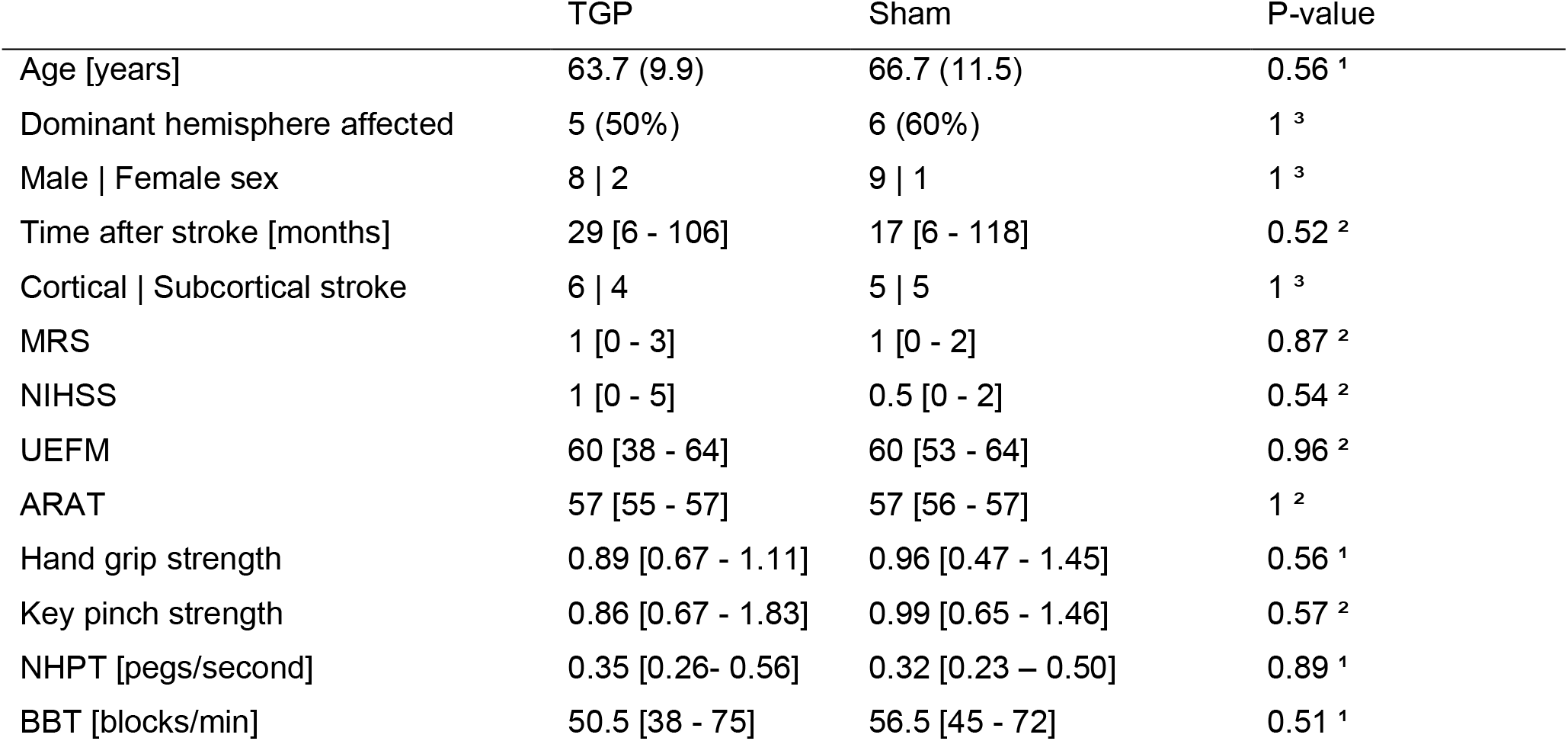
Comparison of clinical characteristics of stroke survivors between stimulation groups. Individuals in the TGP and sham group did not differ significantly in clinical characteristics. Mean (standard deviation) or median [range] values are given per stimulation group. Abbreviations: EHI = Edinburgh Handedness Inventory, NHPT= Nine Hole Peg Test, UEFM = Upper Extremity Fugl-Meyer Assessment, ARAT = Action Research Arm Test, BBT = Box and Block Test, NIHSS = National Institutes of Health Stroke Scale, mRS = modified Rankin Scale. Grip strength values are presented as ratios between the affected and unaffected arm. Uncorrected p-values obtained from: ^1^unpaired t-test, ^2^ Wilcoxon rank-sum test, ^3^ Fisher’s exact test.

**Figure 2:**
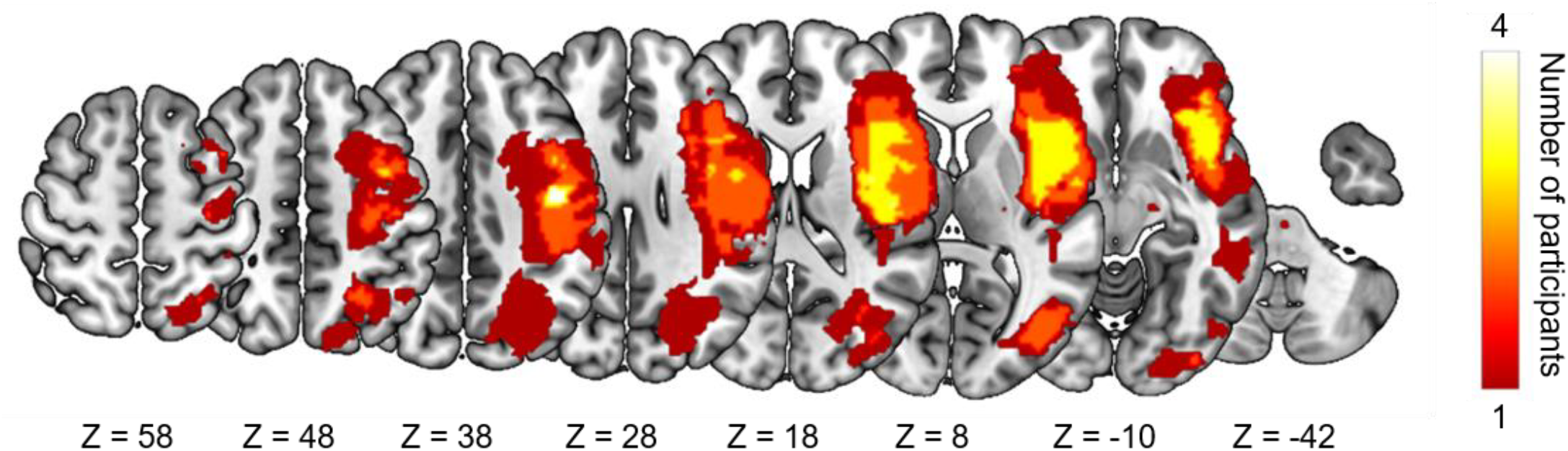
Lesion map. Stroke lesions of the 10 stroke survivors with an available study MRI, overlaid on a T1-weighted image in MNI standard space, including Z-values. The color indicates the number of stroke survivors with lesions at the respective voxel. Right-hemispheric lesions were flipped to the left hemisphere.

### Performance in thumb movement task

On average, young participants made mistakes in 3.3 % of baseline trials and 5.6 % of trials in the stimulation blocks. Outlier removal led to an exclusion of 1.0 % of baseline and 1.4 % of post-baseline trials. Neither the number of mistakes nor outliers differed significantly between stimulation conditions. In the stroke cohort, participants made mistakes in 13.8 % of baseline and 5.3 % of post-baseline trials. 1.3 % of baseline and 1.3 % post-baseline trials were removed as outliers. Stroke survivors made significantly more mistakes during the stimulation blocks when receiving TGP than when receiving sham stimulation (6.9 % vs. 3.7 %, t(18) = 2.4, p = 0.03). There was no difference in the number of mistakes in the baseline block or the number of outliers between the two conditions. For detailed statistics on mistakes and outliers, see Supplementary Table 6-7. To validate the motor skill acquisition task, we assessed whether participants, on average, improved their performance from baseline to the last block. In both cohorts, movement duration decreased significantly (stroke: 0.23 ± 0.19 s (mean ± sd), t(19) = 5.39, p < 0.001; young: 0.18 ± 0.12 s, t(77) = 13.68, p < 0.001).

### Effects of theta-gamma tACS on motor skill acquisition

Our primary hypothesis was that motor skill acquisition would be improved by TGP stimulation compared to TGT and sham stimulation. In healthy participants, we found no differences in motor skill acquisition between TGP (23 ± 9 %) and TGT stimulation (25 ± 11 %; (t(50) = −0.49, p = 0.63) or between TGP and sham stimulation (25 ± 11 %; t(50) = −0.60, p = 0.55, Figure 3A left panel). Thus, motor skill acquisition was not significantly improved by TGP tACS in healthy individuals. We further investigated possible tACS effects on movement duration (Figure 3B, left panel) in a linear mixed-effects model (LME). In healthy participants, we found a significant main effect of block on movement duration (X^2^(1) = 55.09, p < 0.001, 95% CI [-14.5, −9.3]) but no significant main effect of condition (X^2^(2) = 1.67, p = 0.43) nor a condition x block interaction (X^2^(2) = 2.36, p = 0.31). Hence, participants improved their performance over the course of the experiment as expected, but neither TGP nor TGT tACS had a significant effect on overall performance or improvement. In the stroke cohort, contrary to our hypothesis, motor skill acquisition was decreased in the TGP group (12 ± 7 %) compared to the sham group (27 ± 13 %; t(13.6) = −2.71, p = 0.017, Figure 3A right panel).

**Figure 3:**
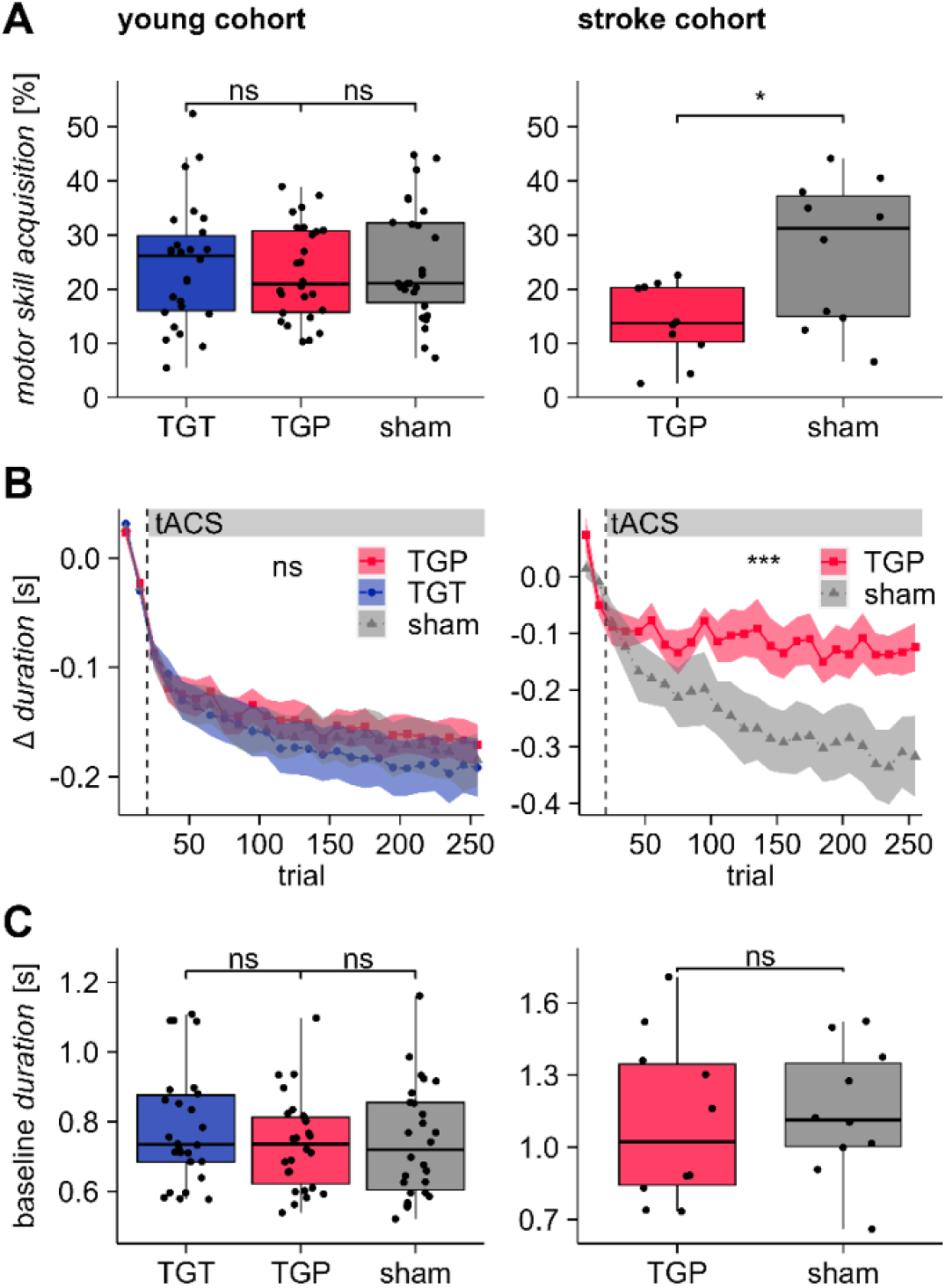
TACS effect on motor skill acquisition. (A) In the young cohort, motor skill acquisition did not differ between TGT, TGP and sham stimulation (left panel), whereas in the stroke cohort, motor skill acquisition was inferior with TGP stimulation compared to sham stimulation (right panel). (B) Time course of mean movement duration per tACS condition relative to the individual baseline. The time course of movement duration did not differ across stimulation conditions in the young cohort (left panel). In the stroke cohort, movement duration showed a greater decrease in the sham condition than in the TGP condition (right panel). The connected data points each represent the mean of 10 consecutive trials. The shaded areas represent the standard error of the mean across individuals. The dashed vertical line marks the end of the baseline and the start of stimulation. (C) Baseline movement duration did not differ significantly across tACS conditions in both the young cohort (left panel) and stroke cohort (right panel). ns: not significant, *: p < 0.05, ***: p < 0.001

Furthermore, in stroke survivors, we found a significant condition x block interaction effect on movement duration (X^2^(1) = 13.5, p < 0.001). Post-hoc analyses revealed that the linear slope of block was significantly steeper in the sham compared to the TGP condition (t(22.2) = 4.16, 95 % CI [13.3, 39.6]). This interaction suggests that TGP stimulation had a detrimental effect on the stroke survivors’ ability to show improvement throughout the task (Figure 3B, right panel). Considering that baseline performance might influence subsequent skill acquisition, we compared movement duration in the baseline block between the tACS conditions (Figure 3C) and found no significant differences between stimulation groups (healthy: TGP vs. TGT; z = −0.76, p = 0.45, TGP vs. sham; t(50) = −0.13, p = 0.89, stroke: TGP vs. sham; t(18) = −0.26, p = 0.80).

### Effects of theta-gamma tACS on acceleration

Motivated by Akkad et al. [4] demonstrating that TGP stimulation enhanced peak thumb acceleration in a ballistic thumb movement task, we conducted an exploratory analysis of peak thumb acceleration in our data (Figure 4A). In the young cohort, we found a significant effect of block (X^2^(1) = 9.62, p = 0.002, 95 % CI [0.13, 0.55]) and stimulation condition (X^2^(2) = 10.96, p = 0.004) on peak acceleration. The interaction of condition x block did not improve the LME (X^2^(2) = 1.63, p = 0.44). The effect of block indicates that healthy participants, in general, increased their peak acceleration over the task. Post-hoc analysis revealed a significantly higher peak acceleration in the TGP condition compared to sham (t(80.2) = 3.08, p = 0.008, 95 % CI [0.60, 4.73]) as well as in the TGT condition compared to sham (t(80.2) = 2.59, p = 0.031, 95 % CI [0.17, 4.27]). There was no significant difference in peak acceleration between TGP and TGT (t(80.2) = 0.53, p = 0.86, 95 % CI [-1.58, 2.48]).

**Figure 4:**
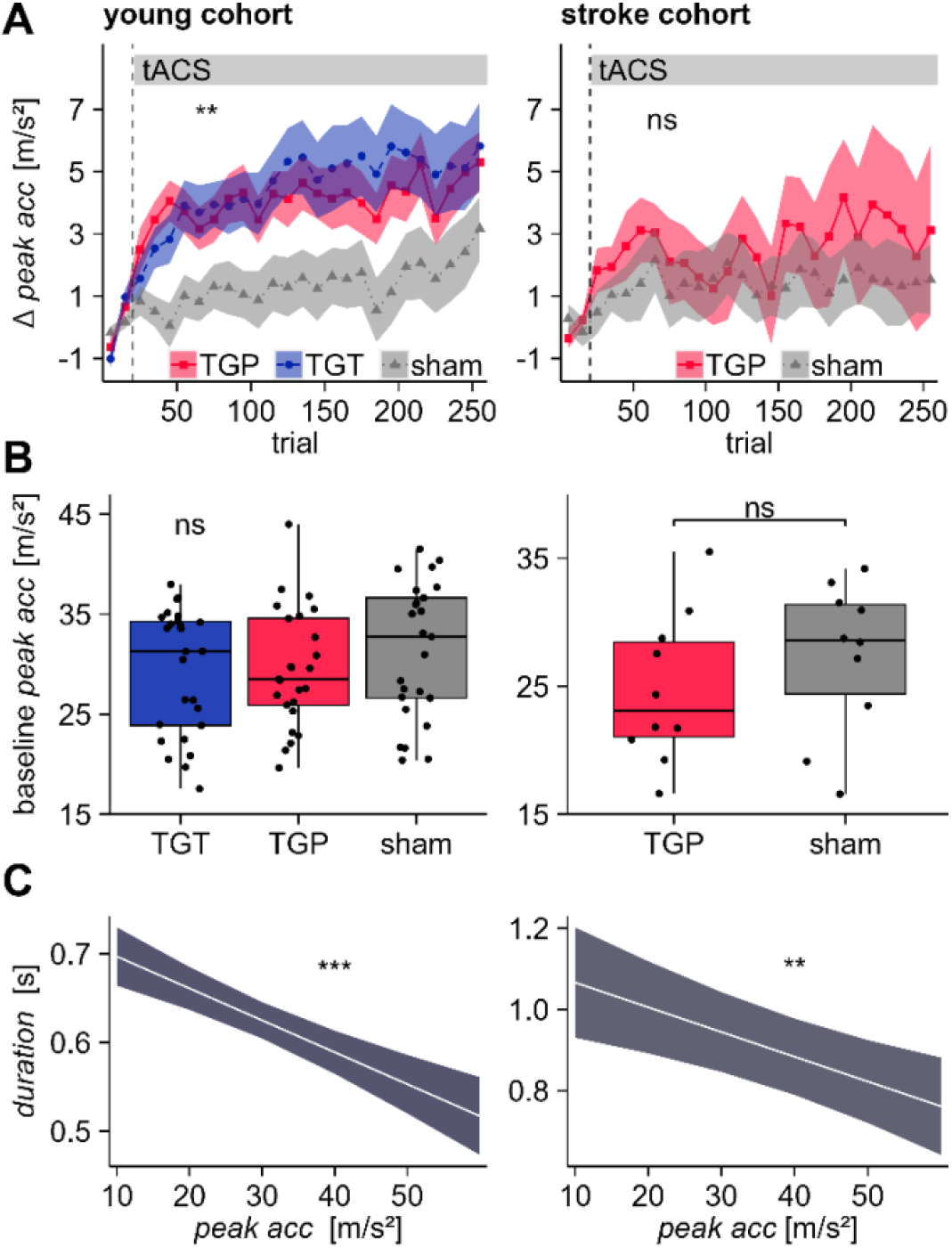
TACS effect on peak acceleration. (A) Both TGP and TGT tACS increase peak acceleration in the young cohort (left panel). In the stroke cohort, peak acceleration did not differ between TGP and sham stimulation (right panel). The connected data points each represent the mean of 10 consecutive trials. The shaded areas represent the standard error of the mean across individuals. The dashed vertical line marks the end of baseline and start of stimulation (B) Baseline peak acceleration did not differ significantly across tACS conditions in both the young cohort (left panel) and stroke cohort (right panel). (C) LME-effect plots of the effect of peak acceleration on movement duration. An increase in peak acceleration is significantly associated with a decrease in movement duration in both cohorts (left panel: young cohort, right panel: stroke cohort). ns: not significant, **: p < 0.01, ***: p < 0.001

In the stroke cohort, most participants showed an increase in peak acceleration over the task (Figure 4A, right panel). Still, we did not find a significant main effect of block (X^2^(1) = 0.33, p = 0.56). Further, there was no significant main effect of condition (X^2^(1) = 0.04, p = 0.84) or significant condition x block interaction (X^2^(1) = 0.13, p = 0.72) either. Peak acceleration during the baseline interval did not differ between stimulation groups, neither in the young nor in the stroke cohort (Figure 4B).

To further understand those results, we analyzed the general relationship between movement duration and peak acceleration. We found that in both cohorts, increased peak acceleration was statistically associated with a shorter movement duration (young: X^2^(1) = 25.03, p < 0.001, 95% CI [-4.9, −2.3]; stroke: X^2^(1) = 9.42, p = 0.002, 95% CI [-9.7, −2.5], Figure 4C).

### Behavioral tACS effects are not significantly related to tACS-induced skin sensations

Behavioral effects of tACS can also be caused by stimulating peripheral nerves on the scalp [41]. Therefore, we applied an anesthetic cream to reduce the activation of peripheral nerves and make tACS more tolerable. Consequently, participants reported only mild to intermediate skin sensations (Figure 5A). Skin sensations showed a not statistically significant trend to be more frequent in the active stimulation groups than in the sham group (young: TGP: 65 %, TGT: 62 %, sham: 42 %, p = 0.22; stroke: TGP: 60 %, sham: 30 %, p = 0.37; uncorrected p-values). There was no significant difference in the frequency of side effects between the cohorts (young: 56 %, stroke: 45 %, p = 0.45).

**Figure 5:**
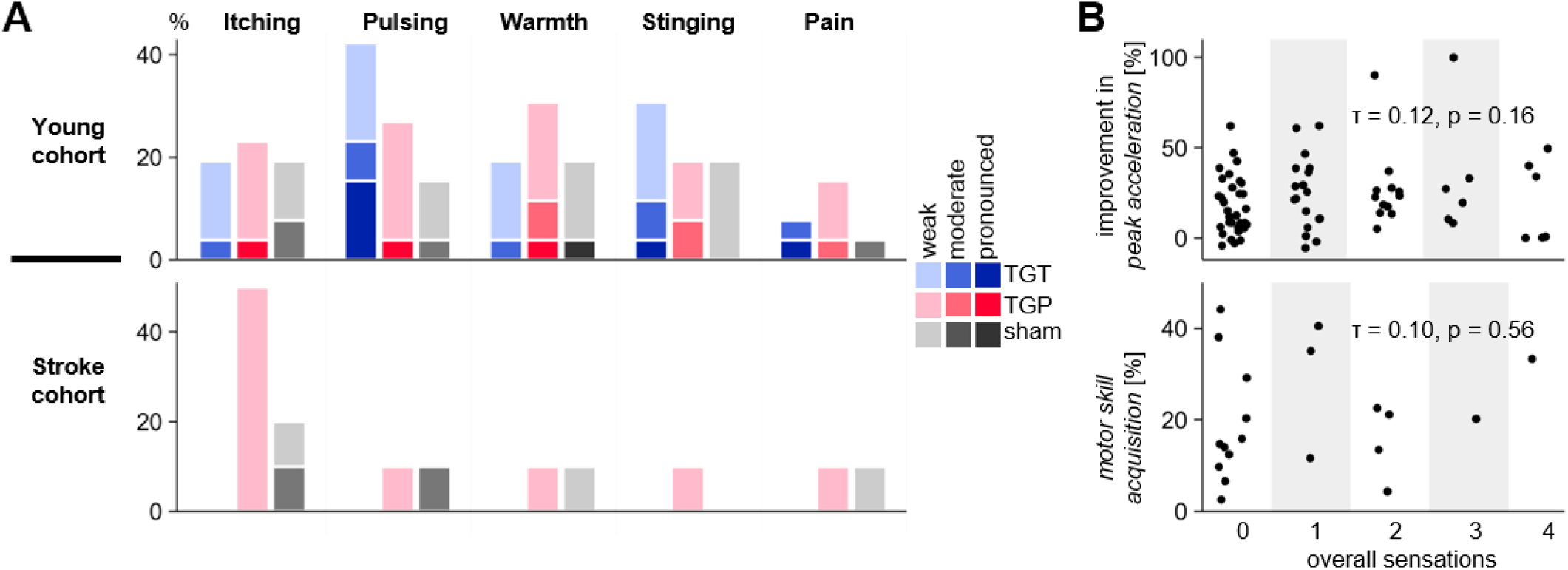
Reported tACS-induced skin sensations (A) Frequency and intensity of skin sensations for different qualities in the young cohort (upper panel) and stroke cohort (lower panel) (B) Top: Depiction of peak acceleration improvement in the young cohort vs. overall skin sensation intensity, bottom: Motor skill acquisition in the stroke cohort vs. overall skin sensation intensity. Kendall correlation coefficient τ and p-values.

If peripheral nerve stimulation played a causal role in the observed motor effects, we would expect motor performance parameters to vary with the intensity of reported skin sensations. However, the correlation between overall skin sensations and motor skill acquisition in the stroke cohort and between overall skin sensations and peak acceleration improvement in the young cohort was not significant (Kendall’s correlation coefficients ≤ 1.2, p > 0.05, Figure 5B). In line with this, motor skill acquisition and peak acceleration improvement did not correlate significantly with the intensity of the single sensation qualities in the young and stroke cohort, respectively (Supplementary Table 2). Skin sensations were significantly more frequent among young female participants (74 %) than young male participants (36 %; p = 0.001, Supplementary Figure 2). As female and male participants were equally distributed across stimulation groups (14 female and 12 male in each group), we do not expect a bias in the group comparisons. In summary, these results suggest a negligible contribution of tACS-induced skin sensations to the motor effects observed in this study. In line with overall low skin sensations, most participants guessed they had received sham stimulation. In both cohorts, the same proportion of participants assumed that they received sham stimulation in all conditions (young: TGP, TGT, sham: 69 %; stroke: TGP: 70 %, sham: 80 %, p = 1). Most participants (58 %) felt unsure of their guess.

### Response to tACS is not biased by clinical characteristics

We found no correlation between motor skill acquisition and stroke survivors’ clinical characteristics (Figure 6, Supplementary Table 3, Spearman correlation coefficients < 0.2, all uncorrected p > 0.4). As the distribution of stroke locations differed slightly between the TGP and sham group, we re-calculated the movement duration LME with matched groups of 8 participants each, with 4 cortical strokes in each group, considering all 375 possible combinations. In an analogous analysis, matching the groups for sex with 8 male, 1 female each, we computed the models for all 18 combinations. All models rendered a significant condition x block interaction (all uncorrected p < 0.01) with a more negative slope, reflecting higher motor skill acquisition in the sham group. We conclude that the distribution of stroke locations and sexes did not relevantly affect the observed tACS effect.

**Figure 6:**
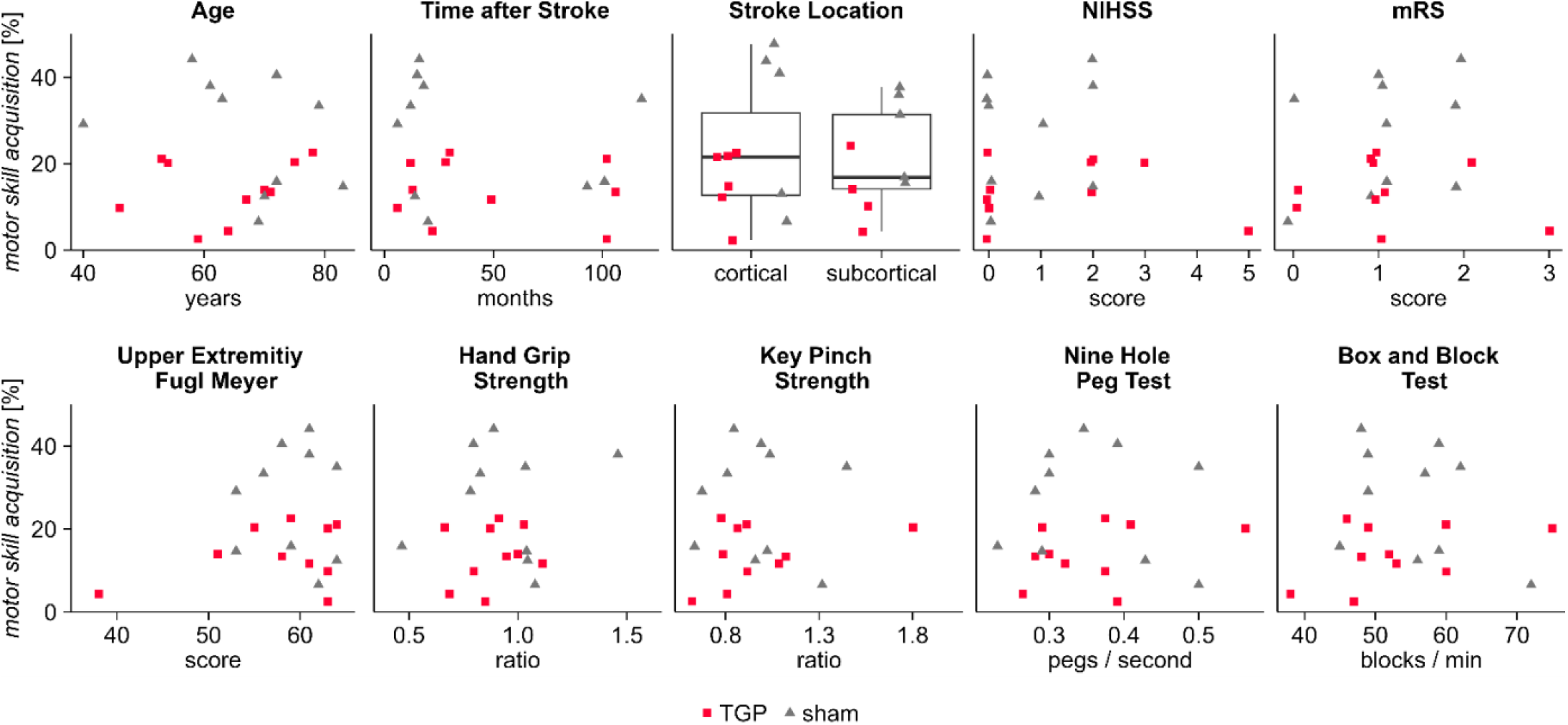
Motor skill acquisition in the stroke cohort displayed against age, clinical scores, motor function scores and stroke lesion characteristics. Grip strength values are presented as ratios between the affected and unaffected arm.

## Discussion

We investigated whether theta-gamma tACS improves the acquisition of a thumb movement skill in a cohort of young, healthy individuals and a cohort of individuals with chronic stroke. TGP tACS deteriorated motor skill acquisition in stroke survivors, while both TGP and TGT tACS did not significantly influence motor skill acquisition in young participants. In an exploratory analysis, we found both TGP and TGT tACS to increase the acceleration of the thumb in the healthy cohort, confirming similar results reported in a previous study [4]. In contrast, we found no significant effect on acceleration levels in the stroke cohort.

These results suggest that theta-gamma tACS does not generally improve motor learning, but can improve specific parameters like the acceleration of movements. Our task differed from the thumb abduction task employed by Akkad et al [4] in terms of the complexity of the movement: the thumb abduction task required pure improvement in acceleration, while our task added coordination skills. This difference in tasks may explain the difference in the primary outcome of the two studies.

To our knowledge, this is the first pre-registered, double-blind, randomized, sham-controlled study investigating the influence of tACS on motor performance in stroke survivors. We found tACS to be highly tolerable in stroke survivors, with a very low level of reported side effects and no adverse effects throughout the whole study. Importantly, successful blinding of the stimulation condition was achieved, as evidenced by very similar probabilities of guessing “sham” or “stimulation” among the conditions.

In opposition to the few existing studies on tACS in stroke survivors, tDCS has been investigated in a large number of stroke cohorts, with very variable outcomes [42–45]. While the mechanism of theta-gamma tACS is unknown so far, one may speculate that the rhythmic depolarization and hyperpolarization of cell membranes can synchronize and desynchronize different networks involved in motor skill acquisition, particularly at theta or gamma frequencies. tACS may thereby produce more specific effects than tDCS.

Specifically, gamma tACS over M1 has prokinetic effects on numerous movement parameters like reaction time [46,47], the amplitude of repetitive movements [48], and the speed and acceleration of force generation [49,50]. It can also improve motor learning [51] and boost motor cortex plasticity in combination with intermittent theta-burst stimulation [52]. Spooner and Wilson [46], however, found that gamma tACS deteriorates movement duration in a sequential finger tapping task. Next to these numerous studies on gamma tACS, there is, to our knowledge, no evidence for effects of theta tACS on motor cortical areas. In addition, we found no significant difference between the effect of TGP and TGT in this study. Thus, it is tempting to speculate that the increased acceleration in young participants primarily relates to gamma tACS. In support of this, we recently found that high-gamma activity scales with movement speed [18] in the same motor task. Future studies may directly compare the effects of gamma tACS with and without theta modulation.

We found divergent effects of theta-gamma tACS on young, healthy participants and stroke survivors and suggest two possible explanations: First, stroke survivors showed lower movement durations and might have had different strategies to improve their movement duration than young participants. Thus, the effects of theta-gamma tACS may be characteristically different due to the differential behavior of the two groups. Second, electrophysiological differences may play a role. The synchronizing effect of tACS is expected to be most prominent when tACS is applied at the resonance frequency of the targeted neuron population [53]. We applied a high-gamma frequency of 75 Hz, the approximate gamma peak frequency for finger movement in young individuals [22,46,54]. However, changes of oscillatory activity [55] and frequency shifts of gamma activity [18,56] occur across the life span. Guerra et al [57] found weaker effects of gamma tACS on the motor cortex in older compared to young participants, hypothesizing a dysfunction or loss of gamma-resonant neurons in older people. A stroke may cause an additional loss of gamma-resonant neurons, leading to different effects of theta-gamma tACS, possibly changing from synchronization to desynchronization or vice versa [58].

Some limitations should be taken into account for this study. First, we did not include electrophysiological recordings like EEG or MEG to determine the frequency or strength of individual theta or gamma oscillations before and after tACS. These parameters may be used to individualize stimulation, to study how stimulation effects depend on these intrinsic frequencies, and to see electrophysiological after-effects of tACS. Thus, we cannot conclude how individual ongoing oscillations may influence the effects of theta-gamma tACS and if tACS changed theta or gamma oscillations. Second, peripheral nerve stimulation may contribute to the effects of tACS, impeding the comparison of sham stimulation to a tACS condition. However, we carefully monitored sensory side effects in a detailed questionnaire. We did not find any statistical relationship between these side effects and the observed tACS effects, making a dominant influence of those side effects unlikely. Third, the stroke cohort is naturally heterogeneous and limited in size. Still, we managed to construct two groups of very high similarity in various clinical and demographic parameters, enabling a comparison of TGP tACS to sham. While the sample size of our stroke cohort is insufficient for subgroup analysis, no significant influence of clinical characteristics on outcome parameters was observed. Fourth, our stroke cohort is restricted to participants in the chronic phase with low impairment who were able to perform the task. For participants with acute stroke or greater impairment, the effects of tACS may differ. Finally, our study does not include a control group of individuals with the age range and lifestyle of stroke survivors. It may be of interest for future studies to see how the effects of theta-gamma tACS depend on age and other factors.

In conclusion, our study confirms that theta-gamma tACS can increase thumb acceleration in healthy young participants. Nevertheless, this increased thumb acceleration may not necessarily translate into improved motor skills in more complex tasks. Most importantly, motor skill acquisition can even be impeded by theta-gamma tACS under pathological conditions such as stroke.

## Data Availability

The data that support the findings of this study are available from the corresponding author, upon reasonable request.

## Acknowledgements

We thank Mareike Gann, Marina Gollmer (née Fiene), Andrew Sharott for helpful discussions, and Jan Feldheim for technical support. Furthermore, we thank all participants who took part in this study.

## Supplementary material

Please see the separate document.

## CRediT author statement

**L. Sophie Grigutsch**: Methodology, Investigation, Software, Formal analysis, Validation, Visualization, Data Curation, Writing - Original draft, Writing - Review & Editing. **Benjamin Haverland**: Methodology, Software, Writing - Review & Editing. **Lena S. Timmsen**: Investigation, Software, Writing - Review & Editing. **Liv Asmussen**: Visualization, Writing - Review & Editing. **Hanna Braaß**: Methodology, Writing - Review & Editing. **Silke Wolf**: Data Curation, Writing - Review & Editing. **The Vinh Luu**: Formal analysis, Visualization, Writing - Review & Editing. **Charlotte J. Stagg**: Conceptualization, Methodology, Writing - Review & Editing. **Robert Schulz**: Validation, Writing - Review & Editing. **Fanny Quandt**: Conceptualization, Methodology, Validation, Writing - Review & Editing, Data Curation, Resources, Supervision, Funding acquisition, Project administration. **Bettina C. Schwab**: Conceptualization, Methodology, Validation, Writing - Original draft, Writing - Review & Editing, Supervision, Funding acquisition, Project administration.

