## Supplementary material for "Differential effects of theta-gamma tACS on motor skill acquisition in young individuals and stroke survivors: a double-blind, randomized, sham-controlled study"

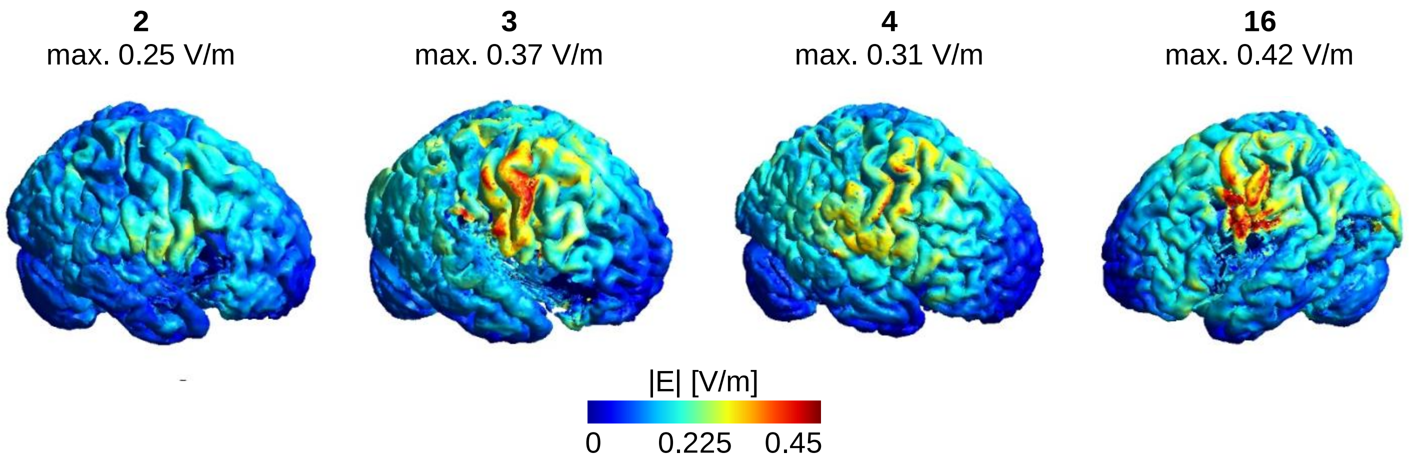


**Supplementary Figure 1: Electric fields of tACS vary in strength and focality.** Electric fields were simulated in SimNIBS for the four participants who received active (TGP) tACS and a head MRI. The participant ID and maximal electric field strength (99$^{th}$ percentile) are given. The lesioned (and thus stimulated) hemisphere is shown.

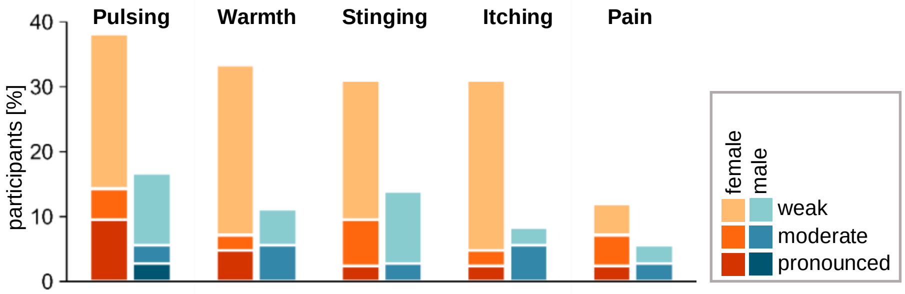


**Supplementary Figure 2: Skin sensations are more frequent among young female than young male participants.**

| Participant ID | Age group | Sex | Lesion side | TAS [months] | Lesion location | Imaging | EHI | mRS | NIHSS | UEFM | ARAT | Grip Strength | Pinch Strength | NHPT [pegs/s] | BBT [blocks/min] |
|---|---|---|---|---|---|---|---|---|---|---|---|---|---|---|---|
| **1** | 71-75 | M | R | 101 | MI, CS | none | 100 | 1 | 0 | 59 | 56 | 0.47 | 0.68 | 0.23 | 45 |
| **2** | 56-60 | M | R | 102 | MI | study MRI | 100 | 1 | 0 | 63 | 57 | 0.85 | 0.67 | 0.39 | 47 |
| **3** | 66-70 | M | R | 13 | MI | study MRI | 100 | 0 | 0 | 51 | 57 | 1.00 | 0.83 | 0.30 | 52 |
| **4** | 46-50 | M | R | 6 | MI, CR | study MRI | 80 | 0 | 0 | 63 | 57 | 0.80 | 0.87 | 0.38 | 60 |
| **5** | 66-70 | M | R | 14 | MI | study MRI[1] | 100 | 1 | 1 | 64 | 57 | 1.04 | 0.98 | 0.43 | 56 |
| **6** | 71-75 | F | R | 15 | CR, PRE | study MRI[1] | 100 | 1 | 0 | 58 | 57 | 0.80 | 1.02 | 0.39 | 59 |
| **7** | 36-40 | M | L | 6 | BG, PLIC | study MRI | 100 | 1 | 1 | 53 | 57 | 0.78 | 0.65 | 0.28 | 49 |
| **8** | 66-70 | M | L | 49 | MI | none | 100 | 1 | 0 | 61 | 57 | 1.11 | 1.08 | 0.32 | 53 |
| **9** | 61-65 | M | L | 18 | MI, PI | clinical CT | 40 | 1 | 2 | 61 | 57 | 1.46 | 1.00 | 0.30 | 49 |
| **10** | 71-75 | M | L | 106 | MI, BG | clinical MRI | 100 | 1 | 2 | 58 | 57 | 0.95 | 1.09 | 0.28 | 48 |
| **11** | 61-65 | M | L | 118 | MO | clinical MRI | 100 | 0 | 0 | 64 | 57 | 1.03 | 1.47 | 0.50 | 62 |
| **12** | 76-80 | M | L | 30 | Pons | clinical MRI | 100 | 1 | 0 | 59 | 57 | 0.91 | 0.80 | 0.38 | 46 |
| **13** | 81-85 | M | L | 93 | Pons | study MRI | 100 | 2 | 2 | 53 | 57 | 1.04 | 1.04 | 0.29 | 59 |
| **14** | 76-80 | M | R | 12 | PLIC | none | 100 | 2 | 0 | 56 | 57 | 0.83 | 0.85 | 0.30 | 57 |
| **15** | 61-65 | F | L | 22 | BG, PLIC | clinical MRI | 100 | 3 | 5 | 38 | 55 | 0.67 | 0.85 | 0.26 | 38 |
| **16** | 51-55 | M | L | 102 | MI | study MRI[1] | -60 | 1 | 2 | 64 | 57 | 1.03 | 0.96 | 0.41 | 60 |
| **17** | 56-60 | M | L | 16 | MI | study MRI | 80 | 2 | 2 | 61 | 57 | 0.89 | 0.89 | 0.35 | 48 |
| **18** | 51-55 | M | R | 12 | MI | clinical MRI | 64 | 1 | 3 | 63 | 57 | 0.87 | 0.83 | 0.56 | 75 |
| **19** | 66-70 | M | L | 20 | PRE,POST | study MRI | 100 | 0 | 0 | 62 | 57 | 1.08 | 1.31 | 0.50 | 72 |
| **20** | 71-75 | F | L | 28 | MI, IC, FO | clinical CT | 80 | 2 | 2 | 55 | 57 | 0.67 | 1.87 | 0.29 | 49 |
| **median\| count** | 68 | 3 F 17 M | 8 R 12 L | 21 | | | 100 | 1 | 0.5 | 60 | 57 | 0.90 | 0.93 | 0.33 | 52.5 |

**Supplementary Table 1: Clinical and demographic data of stroke survivors.**
Abbreviations: TAS = time after stroke. Lesion location: L = left, R = right, BG = basal ganglia, CS = centrum semiovale, CR = corona radiata, FO = frontal operculum, IC = insular cortex, MI = middle cerebral artery infarct, MO = medulla oblongata, PI = posterior cerebral artery infarct, PLIC = posterior limb of the internal capsule, PRE = precentral gyrus; POST = postcentral gyrus; sex: F = female, M = male; clinical tests: EHI = Edinburgh Handedness Inventory, mRS = modified Rankin Scale, NIHSS = National Institutes of Health Stroke Scale, UEFM = Upper Extremity Fugl-Meyer Assessment, ARAT = Action Research Arm Test, NHPT = Nine Hole Peg Test, BBT = Box and Block Test. Grip strength values are presented as ratios between the affected and unaffected arm. [1]MRI data from a previous study in our lab.

| | | Sperman's correlation | | Kendall correlation | |
|---|---|---|---|---|---|
| | | rho | p | tau | p |
| **Young cohort** | Over-all score | 0.16 | 0.16 | 0.12 | 0.16 |
| | Itching | 0.09 | 0.45 | 0.07 | 0.44 |
| | Pulsing | 0.20 | 0.09 | 0.16 | 0.08 |
| | Stinging | 0.05 | 0.65 | 0.04 | 0.68 |
| | Warmth | 0.09 | 0.43 | 0.07 | 0.42 |
| | Pain | 0.12 | 0.30 | 0.10 | 0.28 |
| **Stroke cohort** | Over-all score | 0.14 | 0.54 | 0.10 | 0.56 |
| | Itching | 0.15 | 0.54 | 0.13 | 0.51 |
| | Pulsing | 0.11 | 0.65 | 0.08 | 0.66 |
| | Stinging | -0.22 | 0.35 | -0.18 | 0.34 |
| | Warmth | -0.09 | 0.72 | -0.07 | 0.71 |
| | Pain | 0.17 | 0.46 | 0.15 | 0.45 |

**Supplementary Table 2: The intensity of reported skin sensations is not significantly associated with behavior.** Young cohort: Correlation coefficients of sensation intensities and *peak acceleration improvement*, as tACS significantly modulated *peak acceleration*. Stroke cohort: Correlation coefficients of sensation intensities and *motor skill acquisition,* as *motor skill acquisition* was significantly modulated by tACS. All p-values are uncorrected.

| | Sperman‘s correlation | | Kendall correlation | |
|---|---|---|---|---|
| | rho | p | tau | p |
| Age | 0.05 | 0.84 | 0.10 | 0.56 |
| Time after Stroke | -0.14 | 0.57 | -0.09 | 0.54 |
| mRS | 0.17 | 0.47 | 0.15 | 0.41 |
| NIHSS | 0.07 | 0.77 | 0.06 | 0.75 |
| UEFM | -0.04 | 0.87 | 0 | 1 |
| Nine Hole Peg Test | 0.03 | 0.89 | 0.01 | 0.95 |
| Box and Block Test | 0.08 | 0.72 | 0.04 | 0.79 |
| Hand grip strength ratio | -0.05 | 0.84 | -0.04 | 0.82 |
| Key pinch strength ratio | 0.07 | 0.77 | 0.04 | 0.82 |

**Supplementary Table 3: Clinical characteristics do not significantly correlate with *motor skill acquisition*.** Correlation coefficients and uncorrected p-values.

| **Young cohort - *movement duration* LME** | | | |
|---|---|---|---|
| **Fixed effect** | | **Estimate** | **Standard error** |
| Intercept | | 219.131 | 28.474 |
| Condition: | TGT | 13.838 | 13.589 |
| | Sham | -2.679 | 13.483 |
| Block | | -11.897 | 1.329 |
| Baseline | | 0.568 | 0.036 |

| **Young cohort - *peak acceleration* LME** | | | |
|---|---|---|---|
| **Fixed effect** | | **Estimate** | **Standard error** |
| Intercept | | 6.677 | 1.707 |
| Condition | TGT | -0.449 | 0.817 |
| | Sham | -2.668 | 0.831 |
| Block | | 0.342 | 0.107 |
| Baseline | | 0.872 | 0.054 |

**Supplementary Table 4: Estimates and standard error for fixed effects of LMEs in the young cohort.** Left: *movement duration* LME. Right: *peak acceleration* LME. Reference condition = TGP.

| **Stroke cohort - *movement duration* LME** | | |
|---|---|---|
| **Fixed effect** | **Estimate** | **Standard error** |
| Intercept | 53.201 | 73.919 |
| Sham condition | -12.086 | 38.573 |
| Block | -8.425 | 4.265 |
| Baseline | 0.883 | 0.062 |
| Sham condition x block | -26.459 | 6.028 |

| **Stroke cohort - *peak acceleration* LME** | | |
|---|---|---|
| **Fixed effect** | **Estimate** | **Standard error** |
| Intercept | 7.799 | 1.874 |
| Sham condition | -0.161 | 0.795 |
| Block | 0.131 | 0.227 |
| Baseline | 0.758 | 0.070 |

**Supplementary Table 5: Estimates and standard error for fixed effects of LMEs in the stroke cohort.** Left: *movement duration* LME. Right: *peak acceleration* LME. Reference condition = TGP.

| Mistakes | Total | TGP | TGT | Sham | One-way ANOVA |
|---|---|---|---|---|---|
| Baseline | 3.3 %<br>[0 - 15 %] | 3.9 %<br>[0 - 15 %] | 2.9 %<br>[0 - 15 %] | 3.1 %<br>[0 - 15 %] | F(2,75) = 0.41,<br>p = 0.66 |
| Blocks | 5.6 %<br>[0.4 - 18.8 %] | 4.8 %<br>[0.4 - 13.3 %] | 5.9 %<br>[0.4 - 18.8 %] | 6.0 %<br>[0.4 - 18.3 %] | F(2,75) = 0.46,<br>p = 0.63 |

| Outliers | Total | TGP | TGT | Sham | One-way ANOVA |
|---|---|---|---|---|---|
| Baseline | 1.0 %<br>[0 - 5 %] | 0.4 %<br>[0 - 5 %] | 1.5 %<br>[0 - 5 %] | 1.0 %<br>[0 - 5 %] | F(2,75) = 2.27,<br>p = 0.11 |
| Blocks | 1.4 %<br>[0 – 4.2 %] | 1.5 %<br>[0.4 – 2.9 %] | 1.1 %<br>[0 – 2.5 %] | 1.5 %<br>2.0 [0.4 - 4.2 %] | F(2,75) = 2.4,<br>p = 0.10 |

**Supplementary Table 6: Percentage of mistake trials (top) and outliers (below) in the young cohort.** Comparisons among conditions with one-way ANOVAs rendered no significant differences. Displayed are the means [range].

| Mistakes | Total | TGP | Sham | Comparison |
|---|---|---|---|---|
| Baseline | 13.8 %<br>[0 – 80 %] | 15.5 %<br>[0 – 80 %] | 12.0 %<br>[0 – 25 %] | Wilcoxon rank sum test:<br>z = -0.31, p = 0.76 |
| Blocks | 5.3 %<br>[0.8 - 13.3 %] | 6.9 %,<br>[1.3 - 13.3 %] | 3.7 %<br>[0.8 - 7.5 %] | unpaired t-test:<br>t(18) = 2.4, p = 0.03 |

| Outliers | Total | TGP | Sham | Comparison |
|---|---|---|---|---|
| Baseline | 1.3 %<br>[0 – 5 %] | 1.0 %<br>[0 – 5 %] | 1.5 %<br>[ 0 – 5 %] | Wilcoxon rank sum test:<br>z = -0.45, p = 0.65 |
| Blocks | 1.3%<br>[0.4 - 2.9 %] | 1.4 %<br>2.0 [0.8 – 2.9 %] | 1.1 %<br>[0.4 – 2.1 %] | Wilcoxon rank sum test:<br>z = 0.97, p = 0.33 |

**Supplementary Table 7: Percentage of mistake trials (top) and outliers (below) in the young cohort.** Mean [range]. The comparisons between groups indicate that stroke survivors made significantly more mistakes when receiving TGP tACS compared to sham.
